# Cognitive training and remediation interventions for substance use disorders: A Delphi consensus study

**DOI:** 10.1101/2022.07.28.22278144

**Authors:** Antonio Verdejo-Garcia, Tara Rezapour, Emily Giddens, Arash Khojasteh Zonoozi, Parnian Rafei, Jamie Berry, Alfonso Caracuel, Marc L. Copersino, Matt Field, Eric L. Garland, Valentina Lorenzetti, Leandro Malloy-Diniz, Victoria Manning, Ely M. Marceau, David L. Pennington, Justin Strickland, Reinout Wiers, Rahia Fairhead, Alexandra Anderson, Morris Bell, Wouter J. Boendermaker, Samantha Brooks, Raimondo Bruno, Salvatore Campanella, Janna Cousijn, Miles Cox, Andrew C. Dean, Karen D Ersche, Ingmar Franken, Brett Froeliger, Pedro Gamito, Thomas E. Gladwin, Priscila D. Goncalves, Katrijn Houben, Joanna Jacobus, Andrew Jones, Anne M. Kaag, Johannes Lindenmeyer, Elly McGrath, Talia Nardo, Jorge Oliveira, Charlotte R. Pennington, Kelsey Perrykkad, Hugh Piercy, Claudia I Rupp, Mieke H. J. Schulte, Lindsay M. Squeglia, Petra Staiger, Dan J Stein, Jeff Stein, Maria Stein, William W. Stoops, Mary Sweeney, Hoa Vo, Katie Witkiewitz, Steven P Woods, Richard Yi, Min Zhao, Hamed Ekhtiari

## Abstract

**Background and Aims:** Substance use disorders (SUD) are associated with cognitive deficits that are not always addressed in current treatments, and this hampers recovery. Cognitive training and remediation interventions are well suited to fill the gap for managing cognitive deficits in SUD. We aimed to reach consensus on recommendations for developing and applying these interventions.

**Design:** Delphi approach with two sequential phases: survey development and iterative surveying of experts.

**Setting:** Online study.

**Participants:** During survey development, we engaged a group of 15 experts from a working group of the International Society of Addiction Medicine (Steering Committee). During the surveying process, we engaged a larger pool of experts (n=53) identified via recommendations from the Steering Committee and a systematic review.

**Measurements:** Survey with 67 items covering four key areas of intervention development, i.e., targets, intervention approaches, active ingredients, and modes of delivery.

**Findings:** Across two iterative rounds (98% retention rate), the experts reached a consensus on 50 items including: (i) implicit biases, positive affect, arousal, executive functions, and social processing as key targets of interventions; (ii) cognitive bias modification, contingency management, emotion regulation training, and cognitive remediation as preferred approaches; (iii) practice, feedback, difficulty-titration, bias-modification, goal setting, strategy learning, and meta-awareness as active ingredients; and (iv) both addiction treatment workforce and specialized neuropsychologists facilitating delivery, together with novel digital-based delivery modalities.

**Conclusions:** Expert recommendations on cognitive training and remediation for SUD highlight the relevance of targeting implicit biases, reward, emotion regulation, and higher-order cognitive skills via well-validated intervention approaches qualified with mechanistic techniques and flexible delivery options.

## Introduction

Substance use disorders (SUD) are associated with cognitive deficits that manifest during both active substance use and remission [1–3]. These deficits in executive functions, attention, memory, social processing, and decision-making skills hinder everyday functioning in people with SUD [4–6]. Furthermore, cognitive deficits are associated with difficulties adhering to and benefitting from different SUD treatment programs and settings [7, 8]. Current gold standard treatments for SUD focus on substance use related outcomes, such as drug use reduction or abstinence, often without consideration of cognitive deficits or with the assumption that cognition will recover following successful remission from substance use. However, cognitive deficits can persist even after long-term abstinence and contribute to relapse, reduced quality of life and difficulties reintegrating in society [9, 10]. Furthermore, cognitive deficits are potential obstacles for medication adherence [11] and successful implementation of cognitive behaviour therapies for those with mood, anxiety, and trauma-related comorbidities [12].

Cognitive training and remediation interventions are a logical option to fill the current gap in managing cognitive deficits in SUD [13–15]. These interventions are purpose-built to restore or compensate for cognitive deficits, which may alleviate their impact on daily functioning and improve ability to benefit from SUD treatments, as suggested by demonstrated benefits in other mental health disorders [16]. Moreover, since some cognitive deficits, such as those impacting executive functions and decision-making are not just correlates of SUD but also possibly a core psychopathological mechanism driving compulsive substance use [17, 18], cognitive training and remediation have the potential to become treatments for SUD in and of themselves. Given this premise, it is surprising that this group of interventions have not yet permeated standard care for SUD. This probably relates to the heterogeneity across interventions, and mixed quality of the existing literature [19]. There are numerous small pilot or proof-of-concept trials and comparatively fewer well-powered randomized trials, and there is a wide variety of intervention approaches, with few studies distilling the active ingredients that are purposely driving cognitive and behaviour change [20, 21]. Moreover, most cognitive training and remediation interventions applied in SUD were initially designed for people with other neurological and mental disorders, such as brain injury or schizophrenia, while there are few specific adaptations for people with SUD and addiction treatment programs [22]. Altogether, there are currently very few high-quality, adequately powered and well-structured interventions for improving cognitive functions in SUD. At the same time, cognitive training and remediation for SUD is a growing research area, and both emerging studies and meta-analytic evidence suggests promising benefits for specific approaches [19, 23, 24].

Given the strong rationale for applying cognitive training and remediation interventions in SUD, while acknowledging the heterogeneity and lack of specificity of current approaches, we aimed to reach an expert consensus on recommendations for developing these interventions in the context of SUD. Specifically, we aimed to identify the best strategies for strengthening cognitive functions in people with SUD by surveying experts about the cognitive targets, therapeutic approaches, specific techniques and active mechanisms, and modes of delivery of cognitive training, as well as remediation interventions likely to improve outcomes in the context of SUD treatment. To achieve this, we used a Delphi approach [25, 26] to survey a pool of international experts in the field and reach a broad consensus via iterative consultation.

## Methods

### Pre-registration

The study protocol was pre-registered on the Open Science Framework platform (https://osf.io/xwpes/) on 25/08/2021, prior to commencement of data collection.

### Participants

We engaged two groups of experts during the study: (i) a steering committee (SC), namely, a small and collaborative group of researchers with well-established experience in the field of cognitive training and remediation in SUD who launched the project and interactively developed the initial survey; and (ii) a larger expert panel (EP) who represent the wider community of experts in the field and participated in the surveying. This approach is practical for the procedure of the Delphi study and ensures the quality of the consensus process [27, 28]. Both participants in the SC and EP are co-authors on this paper.

#### Steering Committee (SC)

Following a series of in-person and online meetings within the Neuroscience Interest Group of the International Society of Addiction Medicine (ISAM-NIG) regarding cognitive training and remediation interventions, we established a working group of 15 experts on the topic, which included (in alphabetical order): Jamie Berry, Alfonso Caracuel, Marc Copersino, Hamed Ekhtiari, Matt Field, Eric Garland, Valentina Lorenzetti, Leandro Malloy-Diniz, Victoria Manning, Ely Marceau, David Pennington, Tara Rezapour, Justin Strickland, Antonio Verdejo-García, and Reinout Wiers (henceforth, the SC).

SC members outlined the scope and research questions of the Delphi study, which included four areas pertaining to cognitive training and remediation interventions for SUD: *targets* (i.e., cognitive processes that needed to be addressed), *approaches* (i.e., types of interventions), *techniques / mechanisms* (i.e., active ingredients of the interventions) and *modes of delivery*. Next, the SC designed the original Delphi survey via an interactive process of item development, followed by an iterative process until consensus was reached within the SC on the final set of items. All the comments and revisions during the survey design process were handled by two senior members (AVG and HE). Two assistants (AKZ and EG) facilitated the process and managed all contacts and communications.

#### Expert Panel (EP)

Identification of the expert panel members was based on a systematic literature review (search conducted on 31 July 2021) that yielded 108 cognitive training and remediation studies in SUD. The SC assistants screened the studies to identify key authors in the field to be invited to form the EP. The inclusion criteria regarding entering the EP were as follows: a) appearing among the authors of at least two original publications in the systematic review database; and b) keeping the authorship position of first, last, or corresponding of at least one of the papers. In addition, each of the SC members had the opportunity to nominate a maximum of two other candidates for the EP, based on their own knowledge and networks. The members of the SC were also part of the EP. This process resulted in 86 potential candidates (45 based on the review [note that 5 potential participants identified by the review were already in the SC and 1 was uncontactable], 26 nominated by the SC, and 15 were already members of the SC). We subsequently sent invitation emails to each person, with two reminders sent within two-week intervals in the case of not responding.

### Measures

The original Delphi survey included 67 items. The survey items were those identified by the SC as crucial to interrogate the best-suited set of *targets* (27 items), *approaches* (11 items), *techniques* (10 items), and *modes of delivery* (19 items) for interventions aimed at strengthening cognitive functions in the context of SUD treatment. The main structure of the questions was as follows: “How important do you think [survey item here] is for strengthening cognitive function with the aim of improving the outcomes of addiction treatment?” The items concerning intervention *approaches* included follow-up multiple choice sub-questions for each intervention, in which we inquired about the best timing (“detoxification” [first 2 weeks after cessation/reduction of substance use], “early remission” [first 3 months after cessation/reduction of substance use] or “chronic phase” [more than 3 months after cessation/reduction of substance use]), frequency (“several times per day”, “several times per week”, “once per week” or “monthly”) and duration (“within one month”, “1 to 3 months” or “4 to 12 months”). There was no obligation for the participants to answer all the questions.

### Procedure

The Monash University Human Research Ethics Committee approved the study (Reference: MUHREC #27242) and all participants provided informed consent. The study involved two sequential phases: (i) survey development and revision (conducted by the SC via email), and (ii) survey rating (conducted by the EP via an online survey using Qualtrics software) (**Figure 1**). EP’s responses to the Delphi survey were anonymous, and participants had the option of providing demographic and professional data via an independent survey that was not linked with their Delphi survey responses.

**Figure 1.**
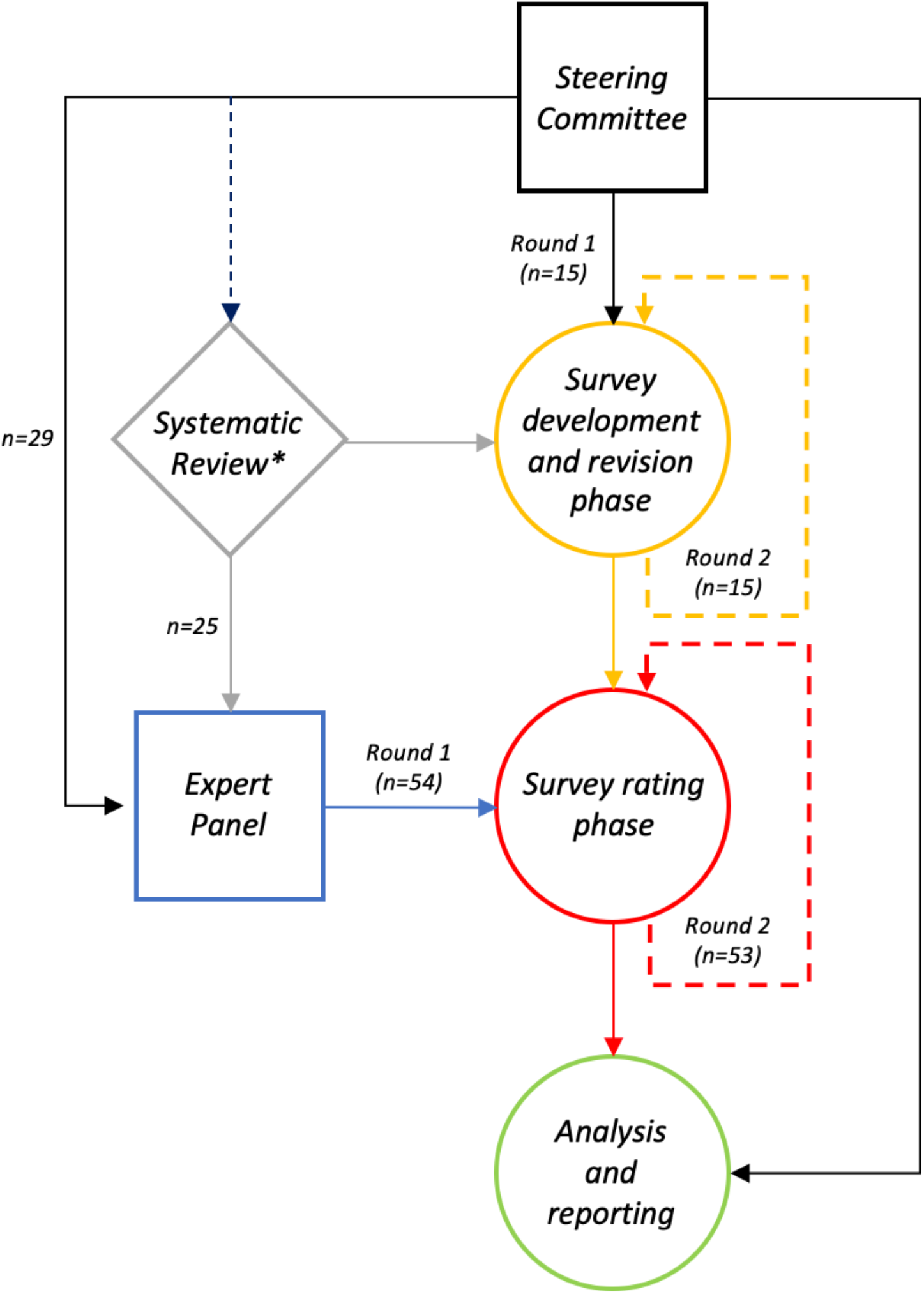
Schema of the study procedure. We had two groups of participants: the Steering Committee (in black) who designed the initial survey draft and participated in all phases of the study, and the Expert Panel (in blue) which includes the Steering Committee along with a broader group of experts in the field derived from a systematic review (in grey) and recommendations by the Steering Committee. The study comprised three main phases including *survey development/revision phase* (in yellow), *survey rating phase* (in red), each happening in two discrete rounds based on reaching consensus and *analysis and reporting phase* (in green). The number of contributors from each source (i.e. Steering Committee [members or nominees] or Systematic Review) is displayed by ‘n=‘.

#### Survey development/revision phase

SC members proposed items to survey the best-suited approaches for improving cognition among people with SUD in the context of SUD treatment, relying on their own experience and knowledge of the evidence base. Upon preparation of the initial draft, all the SC members were asked to add their comments on the survey as a whole and endorse the final version across two rounds of revisions [29]. The final survey was also pilot tested by two senior members of the SC (HE and AVG) to reassure the clarity and coherency of the questions.

A glossary of terms, which contained definitions on every item within the survey, was gathered based on comprehensive literature searches as well as consulting-controlled vocabulary systems of bibliographic databases such as MeSH. This glossary received revisions and final approval by the SC.

#### Survey rating phase

During this phase, the EP rated the finalized version of the survey using their expertise and knowledge [30]. Participants used a five-point Likert scale with the following options: “not important”, “slightly important”, “moderately important”, “very important”, and “essential”. We also provided an “unsure” option. The EP were also invited to suggest new items to be included in the survey.

The consensus threshold was 70%, determined via summation of responses with a score of “moderately important” and above [27, 28]. The procedure was iterative, with experts subsequently surveyed until the greatest possible agreement was reached in a maximum of three rounds, notwithstanding that the same item could not be rated more than two times [25, 26]. The first round included the full set of survey items as agreed by the SC. The second round included: (i) items that reached >50% but less than 70% agreement, and (ii) new items suggested by the experts in the first round. The experts reached consensus after the second round (i.e., 50 items endorsed by >70% of experts), so a third round was not necessary. We have included the two surveys (first and second rounds) as Supplementary Materials.

In order to gauge the degree of diversity in the EP (including the SC), an independent survey, linked via de-identified alphanumeric codes to ensure anonymity, collected socio-demographic and professional information from respondents. Specifically, we collected information regarding age, sex, highest academic degree, country of residence, primary affiliation, primary field of research (psychiatry, psychology, pharmacology, neuroscience, cognitive science, etc.), primary place of work (hospital, university, business, independent research institute, etc.), length of time spent in addiction medicine/science (years), and length of time spent in the field of cognitive rehabilitation in addiction treatment research (years).

### Data Analysis

We computed response percentages for each Delphi survey item and the degree of agreement from participants across the two iterations using IBM SPSS Statistics software version 25. In addition, for those items that were carried forward from the first to the second round, we calculated reliability statistics (Cronbach’s *alpha*) to evaluate the temporal stability of ratings for each item.

## Results

### Participants’ characteristics

Out of 86 original invitations, 59 (68.6%) people responded and 54 (62.7%) completed the first iteration of the survey; the remaining five participants declined because of having moved away from the field (n=3), conflict of interest (n=1), or overcommitment (n=1). Fifty-three (98%) participants completed the second and final iteration of the survey and formed the final expert panel. The expert panel comprised 45% female and 55% male respondents, from geographical locations spanning Africa, the Americas, Asia, Australia, Europe, and the Middle East. They had, on average, 15 years of experience in addiction neuroscience and 10 years of experience in cognitive training and remediation in the context of SUD. They worked across university (80%) and clinical and hospital settings (20%) (**Table 1**).

**Table 1.**
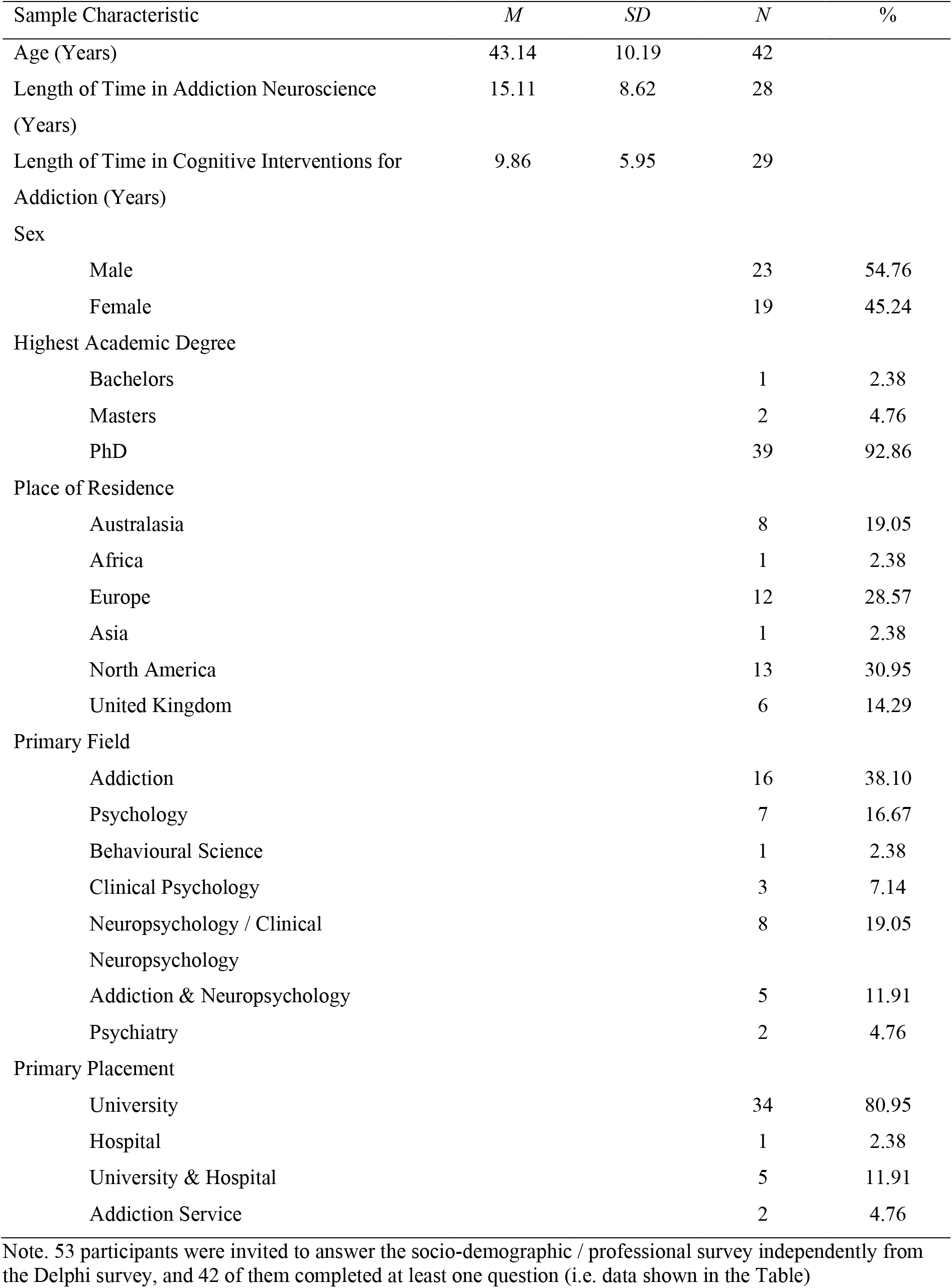
Participants’ socio-demographics and professional characteristics.

### Delphi survey results

**Figure 2** displays the overall flow of the Delphi process, with the number of items from each category endorsed, discarded, or carried forward during subsequent iterations. We achieved consensus after the second iteration with the EP, once all the items had reached pre-established levels of agreement (endorsed) or disagreement (discarded), or had been rated by all experts at least twice without sufficient endorsement [25, 26]. Item reliability across rounds showed adequate consistency (alpha range 0.51-0.75). **Figure 3** shows the pooled experts’ responses (level of agreement) for each item across the two iterations.

**Figure 2.**
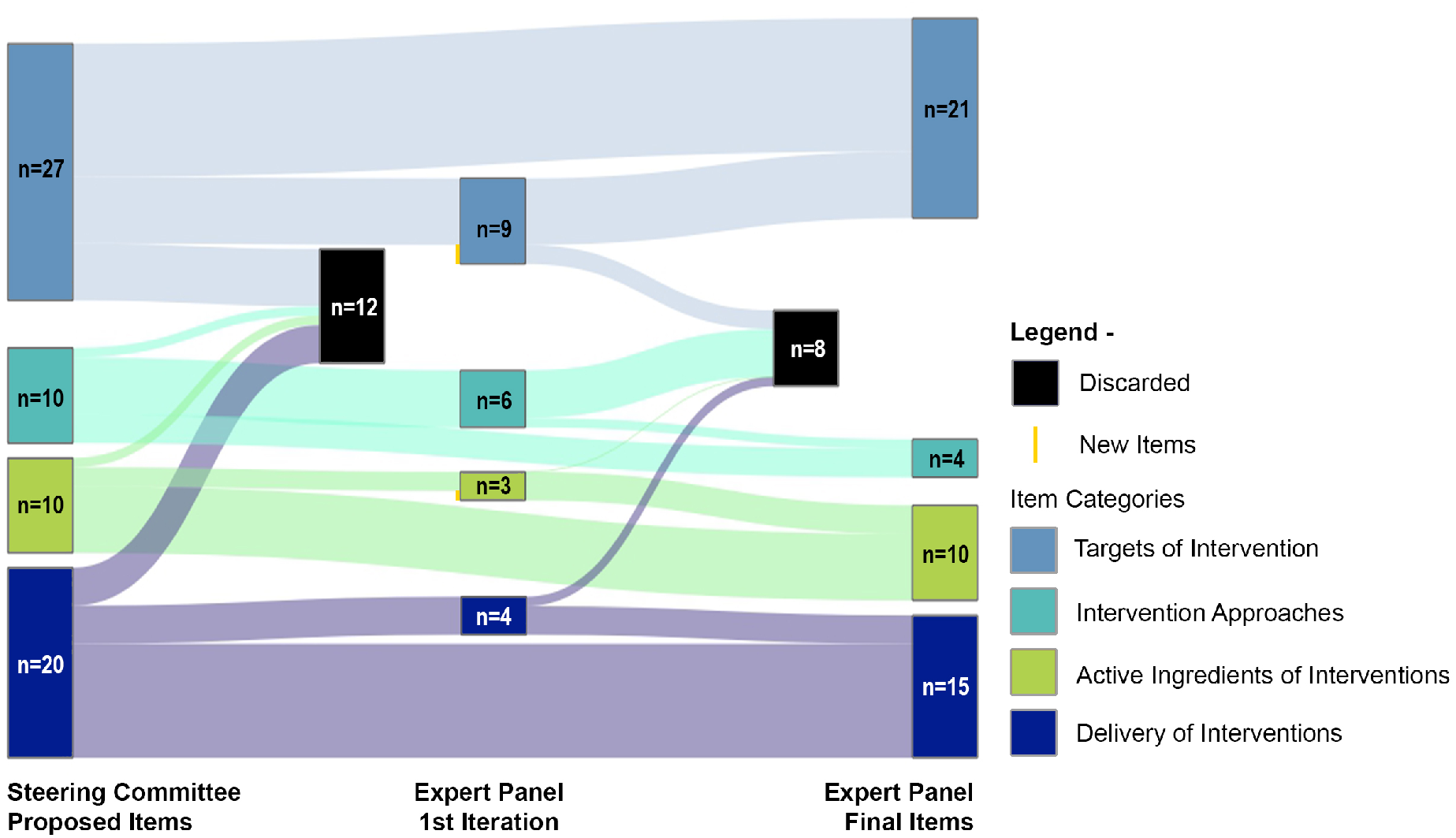
Diagram displaying the flow of the Delphi surveying process. It shows the number of items initially proposed by the Steering Committee for each of the four areas of interest (i.e. targets, approaches, mechanisms or active ingredients and delivery), and how they were subsequently endorsed or discarded by the Expert Panel across two consecutive rounds.

**Figure 3.**
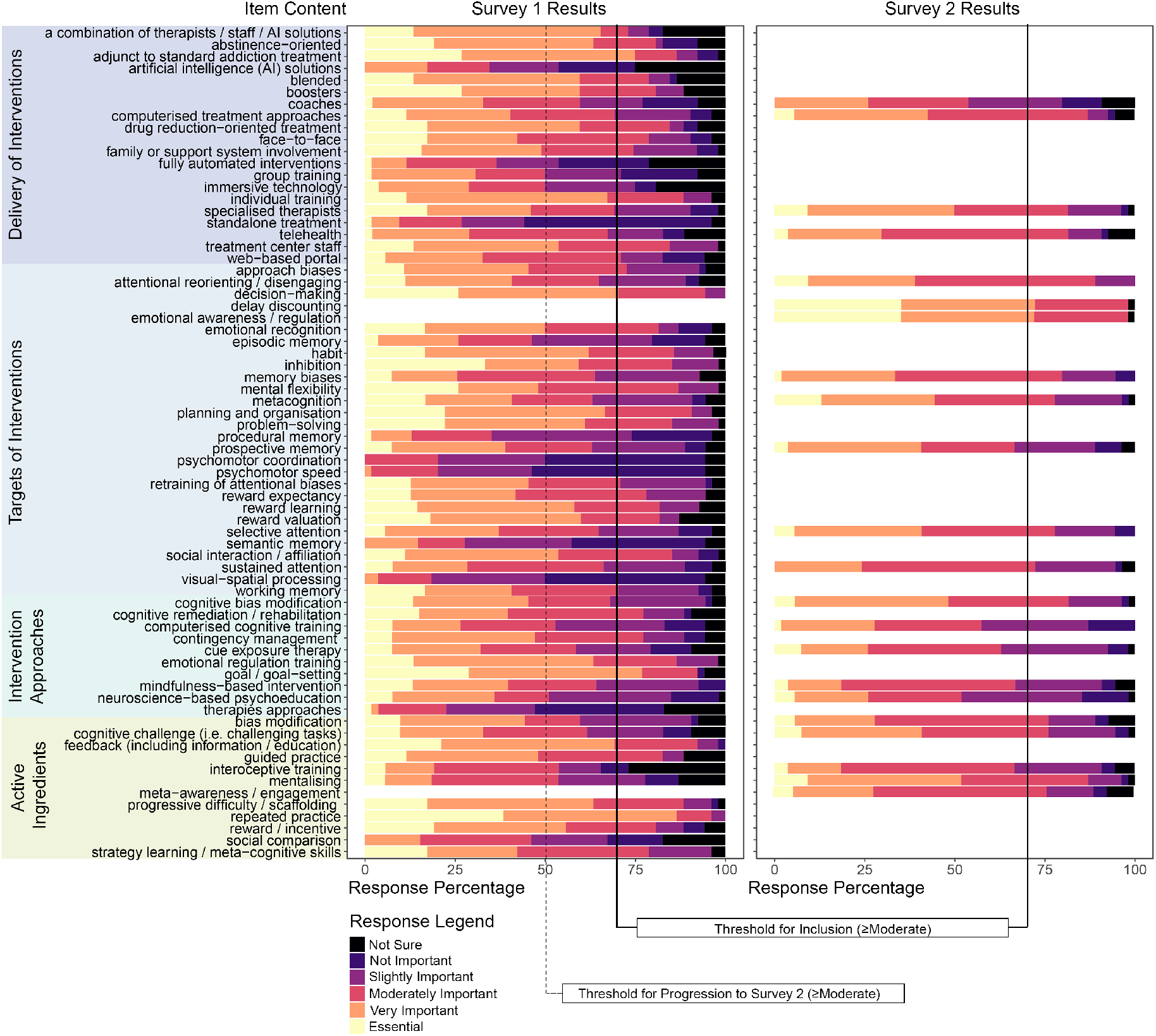
Expert panel participants’ pooled responses to each survey item (i.e. response percentage for each of the Likert scale options), grouped by item category (each of the four areas of interest), across the first and second iterations of the Delphi survey.

Fifty items were endorsed by >70% of the expert panel. **Table 2** enumerates and provides definitions for each of the selected items, organised by category. Definitions were primarily sourced from the American Psychological Association’s Dictionary of Psychology [31] and the National Institute of Mental Health’s Research Domain Criteria (RDoC) Constructs Matrix [32], as well as from specialised literature on cognitive processes (i.e. targets) [33–38], intervention approaches [39–42], techniques / mechanisms [43–45] and modes of delivery [46–48]. In the following subsections, we summarise the results in terms of endorsed and discarded items organised by category.

**Table 2.**
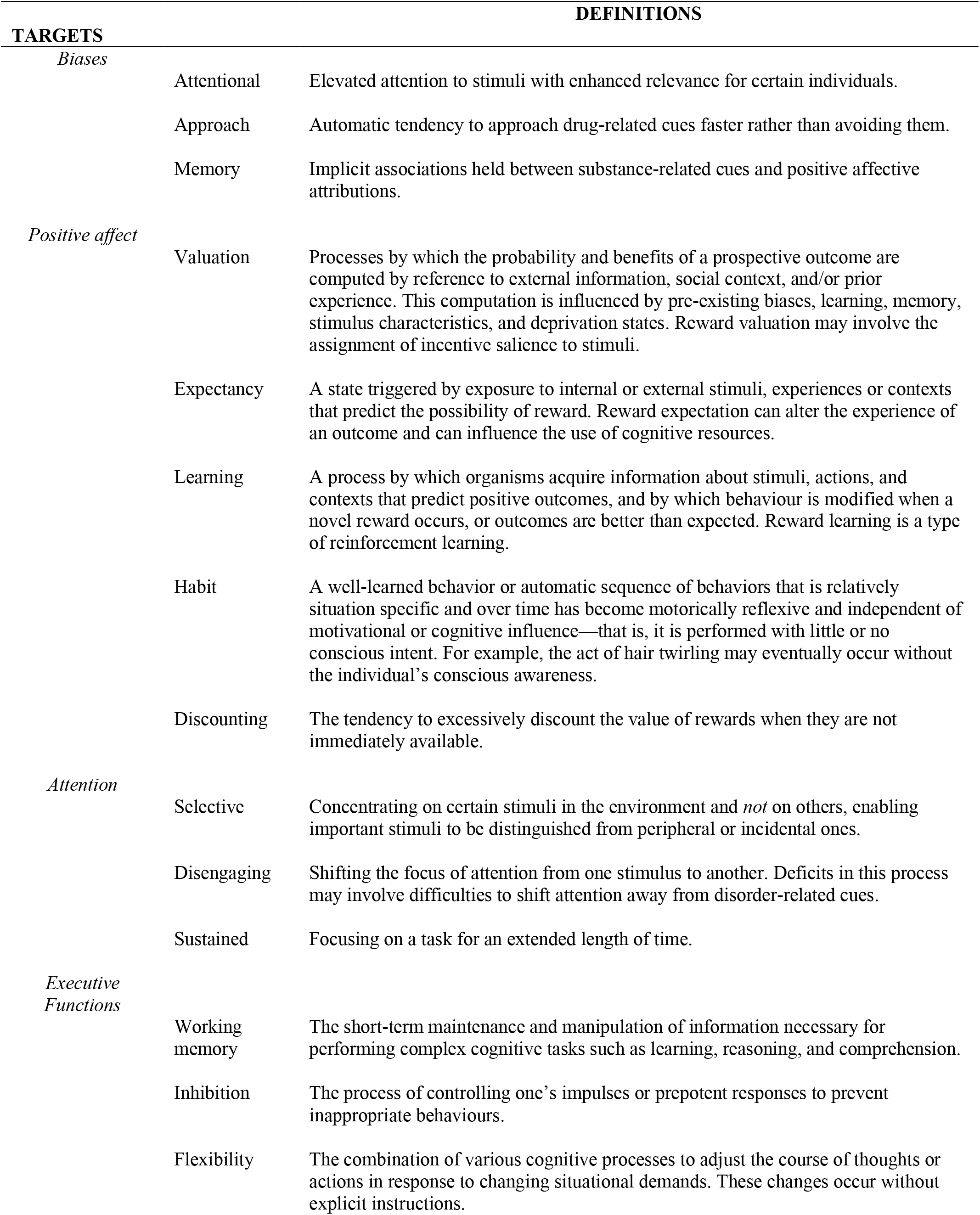
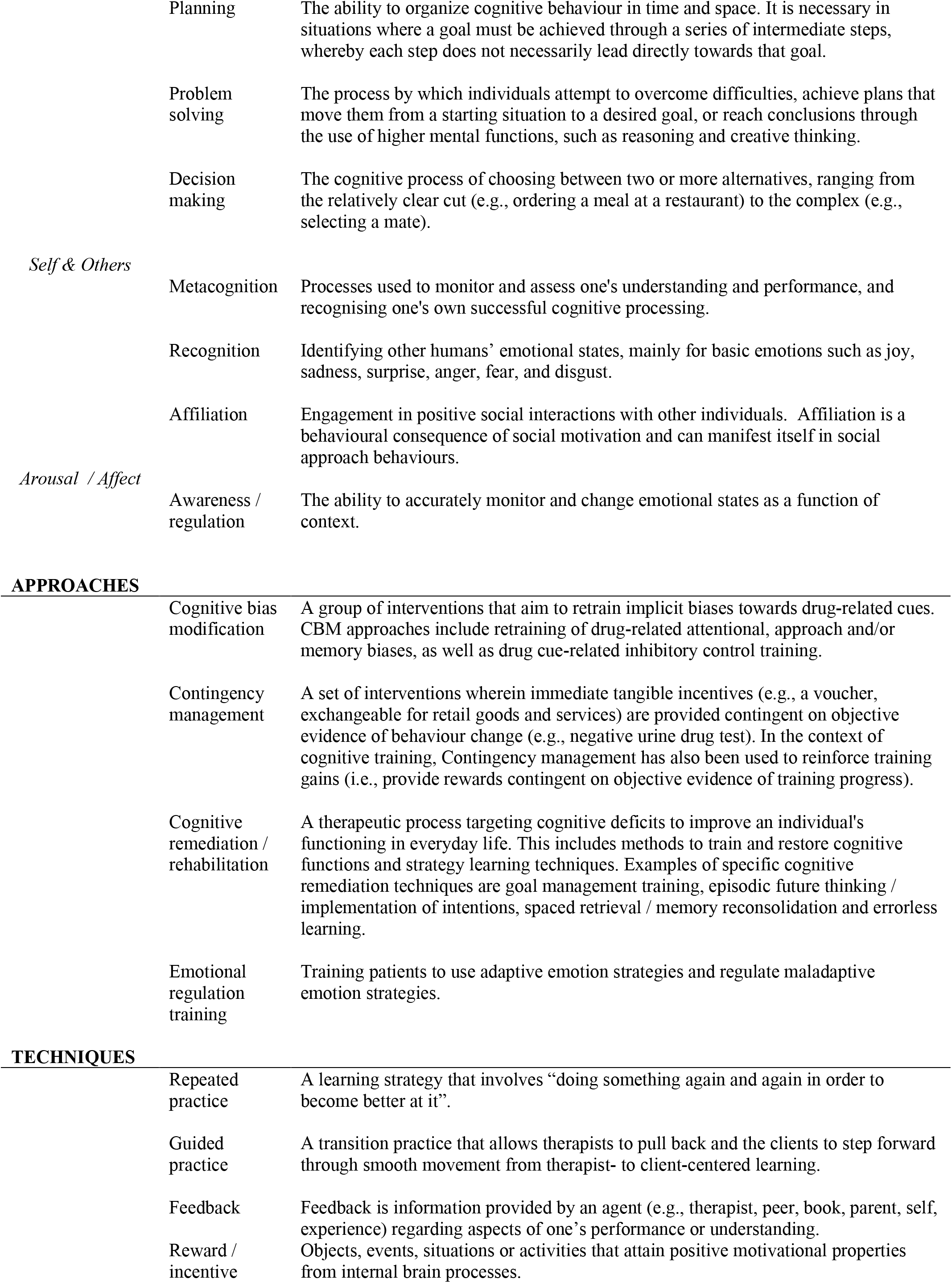
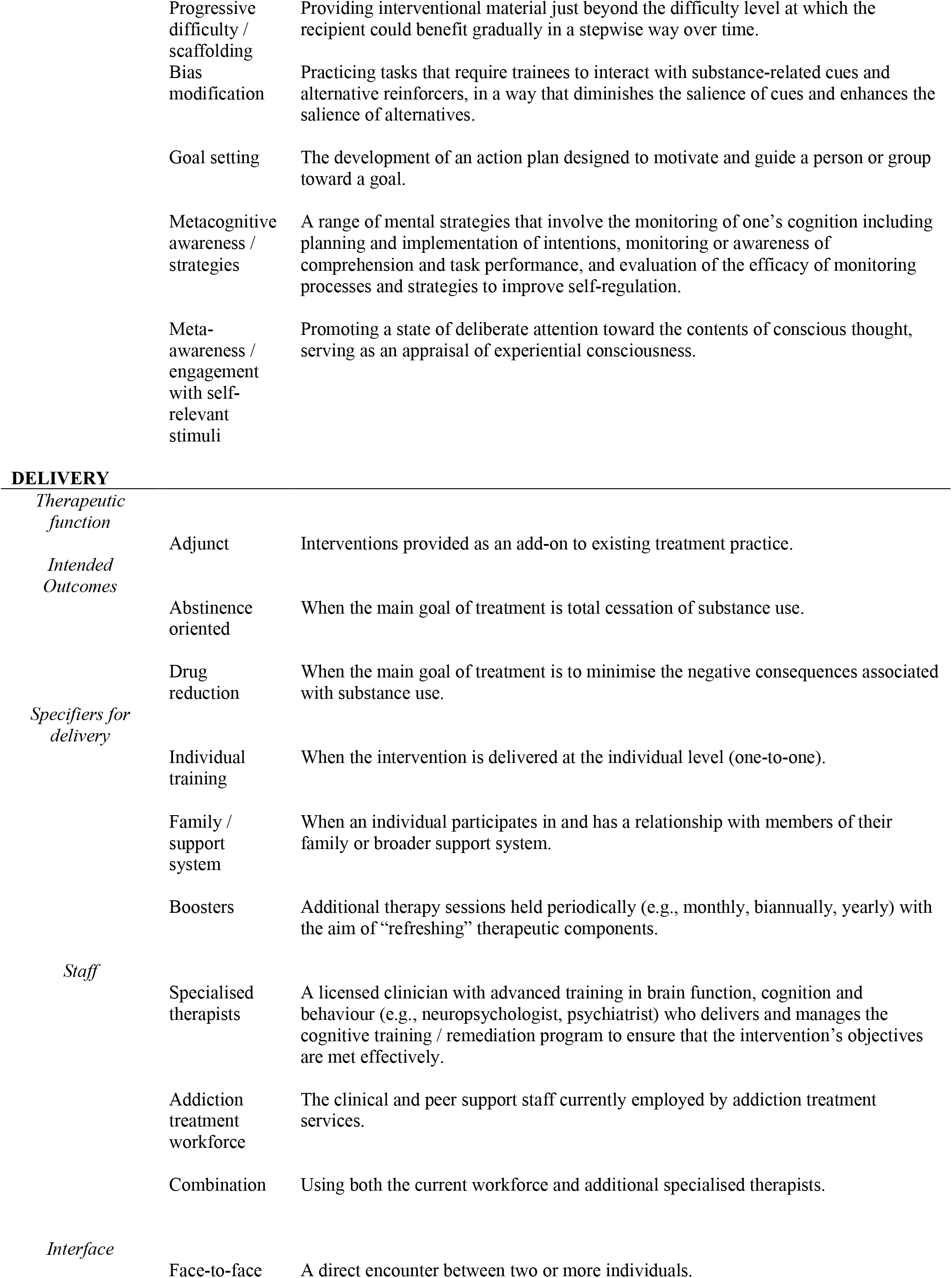
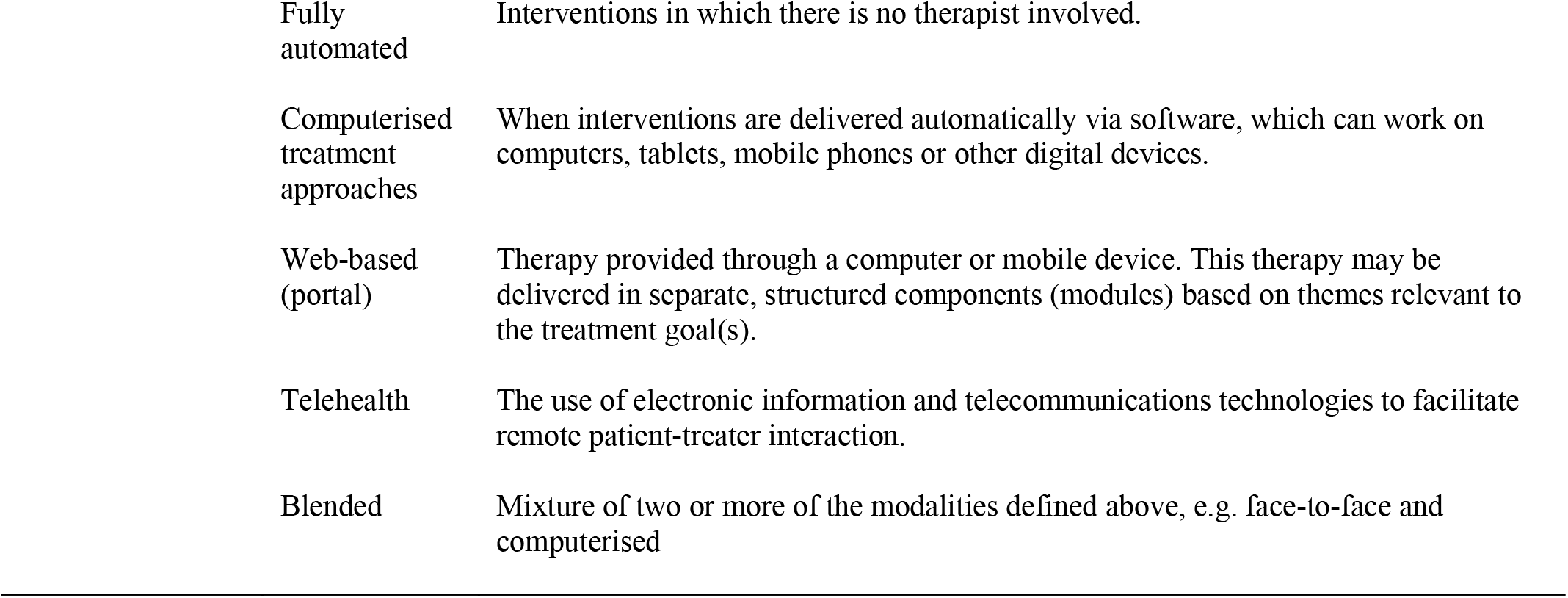
Definitions of final selected items.

#### Targets of intervention

The experts reached consensus on endorsing 21 cognitive processes that should be targeted by cognitive training and remediation interventions (**Table 2**). The selected processes fell into six higher-order systems, namely, cognitive biases, positive affect, arousal and regulatory systems, attention, executive functions, and social processing. The experts discarded eight cognitive processes, including those categorised under perceptual, psychomotor and memory systems, as well as ‘mentalising’ which is part of social systems.

#### Intervention approaches

The experts reached consensus on endorsing four intervention approaches, namely, cognitive bias modification, contingency management, emotion regulation training, and cognitive remediation (**Table 2**). The experts discarded six other approaches, including cue-exposure and aversive therapies, mindfulness and interoceptive trainings, computerised cognitive training, and neuroscience-informed psychoeducation.

For the four interventions endorsed, the majority of experts recommended applying them during early remission (i.e., following detoxification and during the first three months after treatment or self-initiated behaviour change). In terms of frequency, the majority of experts suggested that cognitive bias modification and cognitive remediation should be administered several times per week, whereas for contingency management and emotion regulation training results were more mixed, with preference towards once per week. In terms of duration, the majority of experts suggested that cognitive bias modification and emotion regulation training should be administered over three months, whereas experts suggested longer durations for cognitive remediation and results for contingency management were inconclusive (**Figure 4**).

**Figure 4.**
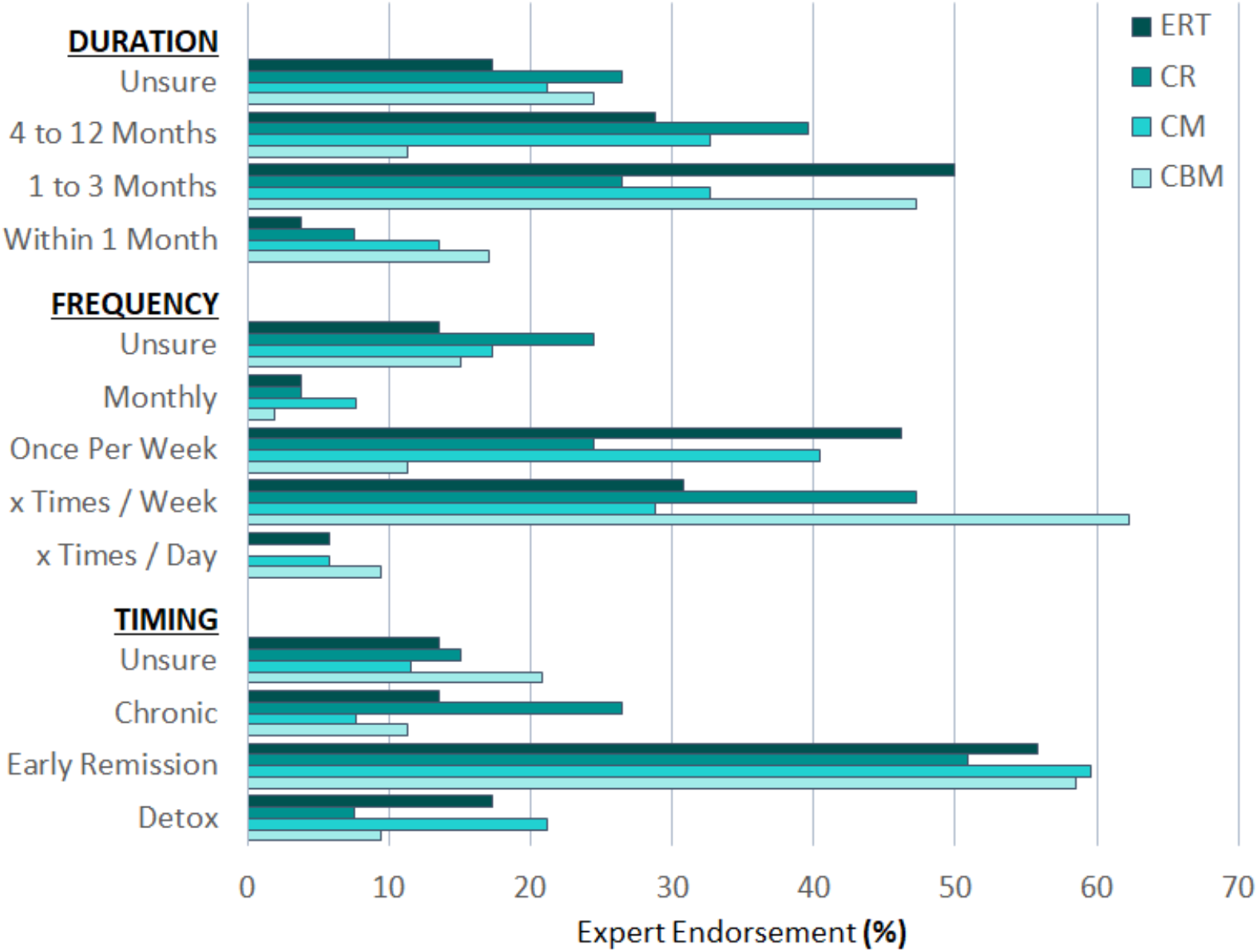
Expert panel participants’ endorsement (percentage responses) of different options regarding timing, frequency, and duration parameters for the selected intervention approaches. *Note*. ERT emotion regulation training, CR cognitive remediation, CM contingency management, CBM cognitive bias modification.

#### Mechanisms / techniques

The experts reached consensus on endorsing 10 different techniques that can be used to initiate and consolidate cognitive training and remediation approaches (**Table 2**). The selected techniques involved different forms of practice (guided and repeated), feedback (information and incentives), titration (progressive difficulty, cognitive challenge), bias modification, goal setting, strategy learning and meta-awareness. Social comparison was the only discarded item.

#### Modes of delivery

The experts reached consensus on endorsing 15 different aspects of intervention delivery (**Table 2**). The experts agreed on considering cognitive training and remediation as an *adjunct* to best-practice interventions. Furthermore, we agreed that cognitive training and remediation could be applied as part of both abstinence-oriented and harm reduction treatment programs. Regarding intervention context, the experts endorsed that cognitive training and remediation interventions should be individually delivered, leverage family support and include boosters to be administered after the active intervention phase. Regarding providers, the experts endorsed involvement of both specialised therapists (e.g., clinical neuropsychologists) and the addiction treatment workforce, as well as combinations of therapists and artificial intelligence. Finally, the experts endorsed several different interfaces for cognitive training / remediation, such as face-to-face, telehealth, web portal, digital / computerised and fully automated, as well as blended approaches. The experts discarded five items, including consideration of cognitive training and remediation as a standalone intervention for addiction treatment, administration in group settings or using immersive technology, and a primary involvement of coaches or artificial intelligence as providers.

## Discussion

This study successfully engaged a pool of 53 experts to provide a consensus on recommended targets, approaches, mechanisms, and delivery of cognitive training and remediation interventions for SUD. The experts agreed on endorsing 50 items across the four different categories (21 targets of intervention, 4 intervention approaches, 10 active ingredients of interventions and 15 aspects of delivery of interventions), and discarded 20 items (8 targets, 6 interventions, 1 mechanism, 5 aspects of delivery). In the following paragraphs, we discuss the logic and implications of each of these four areas of recommendations.

The experts’ selection of targets highlights the key role of cue-related biases, reward, emotion regulation, attention / executive functions, and social processing in the core psychopathology and potential treatment of SUD [20, 49]. This set of processes partly overlaps with those proposed in the Addictions Neuroclinical Assessment (ANA) model (i.e., positive affect, negative affect, executive function) and with the results of a previous Delphi study into neuropsychological assessment for addiction [17, 18]. Our findings go beyond these existing frameworks by highlighting the importance of emotional awareness and regulation, implicit biases, and social processing. Emotional awareness and regulation, which are higher-order (top-down) systems, were emphasized over the basic experience of negative affect, which was discarded for not being well suited for cognitive training and remediation interventions.

Implicit biases and social processes are now included in a hierarchical cognitive model of SUD that strongly aligns with the experts’ view [50]. The selected set of targets also expands and qualifies the scope of previous work by pinpointing, for instance, different types of biases (i.e., approach, attention, memory) and both specific (i.e., working memory, inhibition) and complex (i.e., planning, problem solving) aspects of executive functions. Discarded items (i.e., perceptual, motor and memory processes) are part of the cognitive deficits typically observed in people with SUD. In particular, they include perceptual and motor deficits in people with alcohol use problems and memory problems in those with stimulant use disorders [9]. However, discarded items have a relatively more tenuous relationship with clinical outcomes [7].

The experts endorsed only four (out of the initial ten) intervention approaches. Importantly, there is almost perfect alignment between the endorsed interventions and the selected targets. Cognitive bias modification reduces cue-related biases, contingency management modifies reward processing, emotion regulation training targets emotional awareness and top-down regulation of emotions, and cognitive remediation is the treatment of choice to address attention and executive function deficits in neurological populations [51]. There is no obvious match among selected interventions for social processing targets (i.e., emotion recognition, affiliation) which should be an area of future research, although emotion regulation training partly taps into these processes [52]. The selected interventions have also demonstrated efficacy in the clinical trials literature. There is recent evidence from well-powered randomised controlled trials on the benefits of cognitive bias modification for alcohol abstinence and emotion regulation training for opioid use/misuse reduction [53, 54]. It should be noted, however, that [54] integrated mindfulness with emotion regulation training (i.e., reappraisal and savoring techniques); thus, the robust decrease in opioid misuse observed in this trial may be a function of synergistic interactions between mindfulness and emotion regulation training, as previously proposed [55]. Regarding contingency management, there is a solid evidence base supporting its efficacy for stimulant and opioid use disorders [56, 57]. Although most contingency management trials stemmed from a behavioural economic perspective, rather than being neuroscience-informed, the mechanisms of change of the intervention implicate modifications in neurocognitive processes [58]. In the case of cognitive remediation, there is promising evidence on its ability to improve executive functions from small pilot studies [59], but still limited evidence from well-powered trials, especially about its effects on substance use related outcomes such as craving, use reduction or abstinence [19]. There may also be unknown or unidentified indirect relationships between interventions and targets due to, for example, engagement of parallel or subordinate processes, but those were outside the scope of the study. With regard to non-endorsed interventions, the experts were likely sensitive to the controversies associated with computerised cognitive training [60], which has shown limitations in terms of generalisability of benefits beyond trained tasks [61]. Mindfulness in the absence of explicit emotion regulation training was not supported by the experts despite its appeal among consumers, its efficacy for reducing substance misuse [62, 63], and being an active component of evidence-based interventions for SUD [54]. It is possible that the high degree of heterogeneity in mindfulness interventions, a dearth of studies examining their neurocognitive mechanisms in the context of addiction [64], and the mixed quality of available trials [63] deterred experts from endorsing. Active engagement with cues (as per cognitive bias modification) and cognitive strategies (cognitive remediation) seems to be preferred over the more passive cue-exposure therapies. For other discarded interventions, such as interoceptive training and neuroscience-informed psychoeducation, the lack of research in a still emerging area may have prevented greater support from the experts.

The experts endorsed most of the initially surveyed techniques (ten out of eleven). This suite of techniques can be applied to design specific interventions and intervention regimens, and to monitor consumers’ engagement with active ingredients and its relationship with therapeutic outcomes. In this regard, our selection forms a toolbox that can importantly contribute to the standardisation and systematic evaluation of cognitive training and remediation approaches in the context of SUD. Some of the techniques (e.g., repeated practice) seem better aligned with cognitive training approaches, which seek to restore or reset specific cognitive skills, whereas others such as strategy learning or meta-awareness are usually applied in the context of cognitive remediation [51]. However, an advantage of most of the techniques identified (e.g., guided practice, progressive difficulty and challenge, feedback, bias modification, goal setting) is that they have potential to be transversally applied within several intervention approaches or combinations of approaches. Examples of suitable avenues include the combination of bias modification and feedback techniques in e.g., gamified versions of cognitive bias modification [65], bias modification and strategy learning techniques (e.g., cue-related episodic future thinking) [66] or feedback and strategy learning in combinations of contingency management and goal management [67].

Regarding delivery, the experts supported the role of cognitive training and remediation as an adjunct (rather than a standalone) treatment, which could be integrated in both abstinence-oriented and harm reduction programs. The latter is particularly relevant given increased appraisal of the value of substance use reduction as a therapeutic goal [68]. This amenability to different treatment philosophies, together with endorsement of the existing addiction treatment workforce along with specialized therapists as providers, speaks to the ecological validity and scalability of cognitive training and remediation in the context of standard addiction treatment. Experts also emphasized the need for individual-based delivery. Interestingly, this seemed to be at the cost of discarding group interventions, which include some well-validated approaches that are well accepted by providers and consumers. Through qualitative feedback coming from the experts, we understood that this prioritization of individual-based delivery is due to the perceived need for individualization of the cognitive training and remediation plan, as well as personalization of progress across the intervention. In terms of external support and preferred interfaces, experts embraced multiple sources of support (treatment staff, specialists, family, AI) and multiple / blended interfaces, which can be deemed appropriate depending on particular populations, settings and study designs. The accelerating effect of the current COVID-19 pandemic on remote intervention options likely played a role in leaning experts towards endorsement of telehealth, digital and fully automated approaches. That said, additional large-scale remote intervention trials for telehealth cognitive training and remediation approaches are still needed to ascertain the efficacy of this delivery format. Altogether, the consensus reflects eagerness to embrace the potential of digital health interventions, although caution is needed around the risk for these formats for overpromising and underdelivering [69].

Overall, we leveraged a well-established consensus-reaching method and engaged a diverse group of experts to obtain a comprehensive set of recommendations for the development and implementation of cognitive training and remediation interventions in the context of SUD. We used a two-tiered iterative approach involving both a steering committee and a larger pool of experts, which yielded extremely high retention rates, hence supporting the validity of our findings. The scope of the consensus is unprecedented and our work may pave the way for a new generation of interventions in the SUD treatment arena. There are also limitations to note, such as the modest response rates to our initial invitations (62.7%), which is to some degree expected given our unbiased approach (i.e., based on an independent systematic review) to identify a subsample of the experts. There was also substantial uncertainty (i.e., “unsure” responses) regarding specific survey items, particularly those inquiring about the timing, frequency and duration of interventions, which highlights the need for more empirical research in this area. There was greater representation of invitees from certain geographical locations (e.g., Europe and North America over South America, Africa or Asia) and work settings (i.e., University-based versus clinic / hospital based researchers). Nonetheless, we engaged similar number of male and female participants (45% female) and representation from different locations (i.e., five continents and 13 countries), career stages and expertise (i.e., both cognitive training and rehabilitation experts included) and settings (both basic and clinical researchers and health practitioners were part of our sample), which overall support the diversity of our sample.

## Data Availability

The study protocol was pre-registered on the Open Science Framework platform
(https://osf.io/xwpes/) on 25/08/2021, prior to commencement of data collection.

## Acknowledgement

Prof Alex Baldacchino, for promoting the formation and growth of the Neuroscience Interest Group of the International Society of Addiction Medicine (ISAM-NIG), which was the forum that initiated this Delphi study.

## References

1. Le Berre, A.P., R. Fama, and E.V. Sullivan, Executive Functions, Memory, and Social Cognitive Deficits and Recovery in Chronic Alcoholism: A Critical Review to Inform Future Research. Alcohol Clin Exp Res, 2017. 41(8): p. 1432–1443.

2. Potvin, S., et al., Cocaine and cognition: a systematic quantitative review. J Addict Med, 2014. 8(5): p. 368–76.

3. Verdejo-Garcia, A., G. Garcia-Fernandez, and G. Dom, Cognition and addiction. Dialogues Clin Neurosci, 2019. 21(3): p. 281–290.

4. Hanegraaf, L., et al., A systematic review and meta-analysis of ‘Systems for Social Processes’ in borderline personality and substance use disorders. Neurosci Biobehav Rev, 2021. 127: p. 572–592.

5. Morgan, E.E., et al., Elevated intraindividual variability in methamphetamine dependence is associated with poorer everyday functioning. Psychiatry Res, 2014. 220(1-2): p. 527–34.

6. Zhong, N., et al., The cognitive impairments and psychological wellbeing of methamphetamine dependent patients compared with health controls. Prog Neuropsychopharmacol Biol Psychiatry, 2016. 69: p. 31–7.

7. Dominguez-Salas, S., et al., Impact of general cognition and executive function deficits on addiction treatment outcomes: Systematic review and discussion of neurocognitive pathways. Neurosci Biobehav Rev, 2016. 71: p. 772–801.

8. Stevens, L., et al., Impulsivity as a vulnerability factor for poor addiction treatment outcomes: a review of neurocognitive findings among individuals with substance use disorders. J Subst Abuse Treat, 2014. 47(1): p. 58–72.

9. Fernandez-Serrano, M.J., M. Perez-Garcia, and A. Verdejo-Garcia, What are the specific vs. generalized effects of drugs of abuse on neuropsychological performance? Neurosci Biobehav Rev, 2011. 35(3): p. 377–406.

10. Rubenis, A.J., et al., Impulsivity predicts poorer improvement in quality of life during early treatment for people with methamphetamine dependence. Addiction, 2018. 113(4): p. 668–676.

11. Contardo, C., et al., Relationship of prospective memory to neuropsychological function and antiretroviral adherence. Arch Clin Neuropsychol, 2009. 24(6): p. 547–54.

12. Rezapour, T., et al., NEuro COgnitive REhabilitation for Disease of Addiction (NECOREDA) Program: From Development to Trial. Basic Clin Neurosci, 2015. 6(4): p. 291–8.

13. Nixon, S.J. and B. Lewis, Cognitive training as a component of treatment of alcohol use disorder: A review. Neuropsychology, 2019. 33(6): p. 822–841.

14. Rolland, B., et al., A Patient-Tailored Evidence-Based Approach for Developing Early Neuropsychological Training Programs in Addiction Settings. Neuropsychol Rev, 2019. 29(1): p. 103–115.

15. Verdejo-Garcia, A., Cognitive training for substance use disorders: Neuroscientific mechanisms. Neurosci Biobehav Rev, 2016. 68: p. 270–281.

16. Bowie, C.R., et al., Cognitive remediation for schizophrenia: An expert working group white paper on core techniques. Schizophr Res, 2020. 215: p. 49–53.

17. Kwako, L.E., et al., Addictions Neuroclinical Assessment: A Neuroscience-Based Framework for Addictive Disorders. Biol Psychiatry, 2016. 80(3): p. 179–89.

18. Yucel, M., et al., A transdiagnostic dimensional approach towards a neuropsychological assessment for addiction: an international Delphi consensus study. Addiction, 2019. 114(6): p. 1095–1109.

19. Nardo, T., et al., Cognitive Remediation as an Adjunct Treatment for Substance Use Disorders: A Systematic Review. Neuropsychol Rev, 2022. 32(1): p. 161–191.

20. Verdejo-Garcia, A., et al., A Roadmap for Integrating Neuroscience Into Addiction Treatment: A Consensus of the Neuroscience Interest Group of the International Society of Addiction Medicine. Front Psychiatry, 2019. 10: p. 877.

21. Witkiewitz, K., R.A. Pfund, and J.A. Tucker, Mechanisms of Behavior Change in Substance Use Disorder With and Without Formal Treatment. Annu Rev Clin Psychol, 2022.

22. Rezapour, T., et al., Perspectives on neurocognitive rehabilitation as an adjunct treatment for addictive disorders: From cognitive improvement to relapse prevention. Prog Brain Res, 2016. 224: p. 345–69.

23. Anderson, A.C., et al., Cognitive boosting interventions for impulsivity in addiction: a systematic review and meta-analysis of cognitive training, remediation and pharmacological enhancement. Addiction, 2021. 116(12): p. 3304–3319.

24. Brooks, S.J., et al., Review of the Neural Processes of Working Memory Training: Controlling the Impulse to Throw the Baby Out With the Bathwater. Front Psychiatry, 2020. 11: p. 512761.

25. Yap, M.B., et al., Parenting strategies for reducing the risk of childhood depression and anxiety disorders: A Delphi consensus study. J Affect Disord, 2015. 183: p. 330–8.

26. Yap, M.B., et al., Parenting strategies for reducing the risk of adolescent depression and anxiety disorders: a Delphi consensus study. J Affect Disord, 2014. 156: p. 67–75.

27. Ekhtiari, H., et al., A checklist for assessing the methodological quality of concurrent tES-fMRI studies (ContES checklist): a consensus study and statement. Nat Protoc, 2022. 17(3): p. 596–617.

28. Ekhtiari, H., et al., A methodological checklist for fMRI drug cue reactivity studies: development and expert consensus. Nat Protoc, 2022. 17(3): p. 567–595.

29. Diamond, I.R., et al., Defining consensus: a systematic review recommends methodologic criteria for reporting of Delphi studies. J Clin Epidemiol, 2014. 67(4): p. 401–9.

30. Forsman, A.K., et al., Research priorities for public mental health in Europe: recommendations of the ROAMER project. Eur J Public Health, 2015. 25(2): p. 249–54.

31. VandenBos, G.R., *APA Dictionary of Psychology*. 2nd ed. 2015: American Psychological Association.

32. Health, N.I.o.M. Research Domain Criteria Initiative (RDoC), RDoC Constructs Matrix. 2022 [cited 2022 20-06]; Available from: https://www.nimh.nih.gov/research/research-funded-by-nimh/rdoc/constructs/rdoc-matrix.

33. Stacy, A.W. and R.W. Wiers, Implicit cognition and addiction: a tool for explaining paradoxical behavior. Annu Rev Clin Psychol, 2010. 6: p. 551–75.

34. Owen, A.M., Cognitive planning in humans: neuropsychological, neuroanatomical and neuropharmacological perspectives. Prog Neurobiol, 1997. 53(4): p. 431–50.

35. Fineberg, N.A., et al., New developments in human neurocognition: clinical, genetic, and brain imaging correlates of impulsivity and compulsivity. CNS Spectr, 2014. 19(1): p. 69–89.

36. Fleming, S.M. and H.C. Lau, How to measure metacognition. Front Hum Neurosci, 2014. 8: p. 443.

37. Tracy, J.L. and R.W. Robins, The automaticity of emotion recognition. Emotion, 2008. 8(1): p. 81–95.

38. Berg, E.A., A simple objective technique for measuring flexibility in thinking. J Gen Psychol, 1948. 39: p. 15–22.

39. Gladwin, T.E., C.E. Wiers, and R.W. Wiers, Cognitive neuroscience of cognitive retraining for addiction medicine: From mediating mechanisms to questions of efficacy. Prog Brain Res, 2016. 224: p. 323–44.

40. Kiluk, B.D., et al., Performance-Based Contingency Management in Cognitive Remediation Training: A Pilot Study. J Subst Abuse Treat, 2017. 72: p. 80–88.

41. Barlati, S., et al., Cognitive remediation in schizophrenia: current status and future perspectives. Schizophr Res Treatment, 2013. 2013: p. 156084.

42. Te Brinke, L.W., et al., A cognitive versus behavioral approach to emotion regulation training for externalizing behavior problems in adolescence: Study protocol of a randomized controlled trial. BMC Psychol, 2018. 6(1): p. 49.

43. Jonides, J., How does practice makes perfect? Nat Neurosci, 2004. 7(1): p. 10–1.

44. Wisniewski, B., K. Zierer, and J. Hattie, The Power of Feedback Revisited: A Meta-Analysis of Educational Feedback Research. Front Psychol, 2019. 10: p. 3087.

45. Schultz, W., Behavioral theories and the neurophysiology of reward. Annu Rev Psychol, 2006. 57: p. 87–115.

46. Schlauch, R.C., et al., The moderating effect of family involvement on substance use risk factors in adolescents with severe emotional and behavioral challenges. Addict Behav, 2013. 38(7): p. 2333–42.

47. Fairburn, C.G. and Z. Cooper, Therapist competence, therapy quality, and therapist training. Behav Res Ther, 2011. 49(6-7): p. 373–8.

48. Kumar, V., et al., The Effectiveness of Internet-Based Cognitive Behavioral Therapy in Treatment of Psychiatric Disorders. Cureus, 2017. 9(8): p. e1626.

49. Verdejo-Garcia, A., Neuroclinical Assessment of Addiction Needs to Incorporate Decision-Making Measures and Ecological Validity. Biol Psychiatry, 2017. 81(7): p. e53–e54.

50. Ramey, T. and P.S. Regier, Cognitive impairment in substance use disorders. CNS Spectr, 2019. 24(1): p. 102–113.

51. Cicerone, K.D., et al., Evidence-Based Cognitive Rehabilitation: Systematic Review of the Literature From 2009 Through 2014. Arch Phys Med Rehabil, 2019. 100(8): p. 1515–1533.

52. Nandrino, J.L., et al., Training emotion regulation processes in alcohol-abstinent individuals: A pilot study. Addict Behav, 2021. 114: p. 106652.

53. Manning, V., et al., Effect of Cognitive Bias Modification on Early Relapse Among Adults Undergoing Inpatient Alcohol Withdrawal Treatment: A Randomized Clinical Trial. JAMA Psychiatry, 2021. 78(2): p. 133–140.

54. Garland, E.L., et al., Mindfulness-Oriented Recovery Enhancement vs Supportive Group Therapy for Co-occurring Opioid Misuse and Chronic Pain in Primary Care: A Randomized Clinical Trial. JAMA Intern Med, 2022.

55. Garland, E.L., Restructuring reward processing with Mindfulness-Oriented Recovery Enhancement: novel therapeutic mechanisms to remediate hedonic dysregulation in addiction, stress, and pain. Ann N Y Acad Sci, 2016. 1373(1): p. 25–37.

56. Bolivar, H.A., et al., Contingency Management for Patients Receiving Medication for Opioid Use Disorder: A Systematic Review and Meta-analysis. JAMA Psychiatry, 2021. 78(10): p. 1092–1102.

57. Minozzi, S., et al., Psychosocial interventions for psychostimulant misuse. Cochrane Database Syst Rev, 2016. 9: p. CD011866.

58. Lake, M.T., et al., Decision-Making by Patients With Methamphetamine Use Disorder Receiving Contingency Management Treatment: Magnitude and Frequency Effects. Front Psychiatry, 2020. 11: p. 22.

59. Dubuson, M., et al., Transcranial direct current stimulation combined with alcohol cue inhibitory control training reduces the risk of early alcohol relapse: A randomized placebo-controlled clinical trial. Brain Stimul, 2021. 14(6): p. 1531–1543.

60. Simons, D.J., et al., Do “Brain-Training” Programs Work? Psychol Sci Public Interest, 2016. 17(3): p. 103–186.

61. Caetano, T., et al., Cognitive Training Effectiveness on Memory, Executive Functioning, and Processing Speed in Individuals With Substance Use Disorders: A Systematic Review. Front Psychol, 2021. 12: p. 730165.

62. Li, W., et al., Mindfulness treatment for substance misuse: A systematic review and meta-analysis. J Subst Abuse Treat, 2017. 75: p. 62–96.

63. Korecki, J.R., et al., Mindfulness-based programs for substance use disorders: a systematic review of manualized treatments. Subst Abuse Treat Prev Policy, 2020. 15(1): p. 51.

64. Garland, E.L., et al., Mindfulness-Oriented Recovery Enhancement remediates hedonic dysregulation in opioid users: Neural and affective evidence of target engagement. Sci Adv, 2019. 5(10): p. eaax1569.

65. Manning, V., et al., A Personalized Approach Bias Modification Smartphone App (“SWiPE”) to Reduce Alcohol Use: Open-Label Feasibility, Acceptability, and Preliminary Effectiveness Study. JMIR Mhealth Uhealth, 2021. 9(12): p. e31353.

66. Rafei, P., et al., Imagining the Future to Reshape the Past: A Path to Combine Cue Extinction and Memory Reconsolidation With Episodic Foresight for Addiction Treatment. Front Psychiatry, 2021. 12: p. 692645.

67. Verdejo-Garcia, A., Garcia-Fernandez, G., *Synergistic opportunities in combined interventions for addiction treatment*, in *Cognition and Addiction: A Researcher’s Guide from Mechanisms towards Interventions*, A. Verdejo-Garcia, Editor. 2019, Academic Press: San Diego, USA. p. 405–408.

68. Paquette, C.E., S.B. Daughters, and K. Witkiewitz, Expanding the continuum of substance use disorder treatment: Nonabstinence approaches. Clin Psychol Rev, 2022. 91: p. 102110.

69. Goldberg, S.B., et al., Mobile phone-based interventions for mental health: A systematic meta-review of 14 meta-analyses of randomized controlled trials. PLOS Digit Health, 2022. 1(1).

